# Key Points of Clinical and CT Imaging Features of 2019 Novel Coronavirus (2019-nCoV) Imported Pneumonia Based On 21 Cases Analysis

**DOI:** 10.1101/2020.03.03.20030775

**Authors:** Z.F. Xu, W.X. Wu, Y.B. Jin, A.Z. Pan

## Abstract

**Background and Objective:** WHO Director-General declared that the 2019-nCoV outbreak constitutes a Public Health Emergency of International Concern,and the outbreak is still on-going.Chest CT had been a key component of the diagnostic workup for patients with suspected infection. In this retrospective study, we attempt to summarize and analyze the chest CT features of 2019-nCov infections, and to identify the typical features to improved the diagnostic accuracy of new coronavirus pneumonia (NCP).

**Methods:** Chest CT scans and Clinical data of 21 patients confirmed NCP in our hospital were enrolled.These patients were divided into mild and sever group according to clinical manifestations described by the 6th clinical practice guideline of NCP in China. Main clinical and chest CT features were analyzed and identify.

**Results:** Fever (85.7%) and cough (80.9%) were the two main symptoms of NCP patients.More significantly higher incidence (85.7%) of shortness of breath in the severe cases. Multiple lesions in both lungs and with incidence of GGO(100%),vascular enlargement (76.5%) and cobblestone/reticular pattern(70.6%) were the major feature.The incidence of consolidation, mixed pattern and vascular enlargement features were up to 100% in the severe group, significantly higher than that of patients in mild group. In addition, the incidence of air-bronchogram, dilated bronchi with thickened wall and fibrosis in the severe group was significantly higher than that in the mild group.

**Conclusions:** Fever and cough are the typical clinical features of NCP patients, and chest CT mainly manifested as multiple lesions in both lungs, often accompanied by GGO, vascular enlargement and cobblestone/reticular pattern.Changes in these main CT features can indicate development of the disease

**Summary:** 2019 novel Coronavirus (2019-nCov) had typical clinical manifestations (fever and cough), and presented with characteristic chest CT imaging features (multiple lesions in both lungs, often accompanied by GGO, vascular enlargement and cobblestone/reticular pattern), which are helpful to the radiologist in the early detection and diagnosis of this emerging global health emergency. In addition, changes in these main CT features can indicate development of the disease.

**Highlights:**

1. Fever (85.7%) and cough (80.9%) were the two main symptoms of NCP patients.The incidence of shortness of breath was 85.7% in the severe cases, significantly higher than 21.4% in the mild cases.
2. Multiple lesions in both lungs and with incidence of GGO (100%), vascular enlargement (76.5%) and cobblestone/reticular pattern (70.6%) were the major features of NCP patients. 85.7% of cases in serve group displayed 4-5 lobes were involved simultaneously.
3. Changes in these main CT imaging features can indicate development of the disease. About 19.1% of patients (4 of 21) presented with a normal CT.

## 1. Introduction

The atypical pneumonia case, caused by a novel coronavirus (2019-nCov), was first reported and confirmed in Wuhan, China in December 31, 2019 (Pneumonia of unknown cause-China,2020)[1]. Initially confined to Wuhan, the infection has spread elsewhere, sporadic cases exported from Wuhan were reported in many foreign countries.On January 30, the WHO Director-General declared that the 2019-nCov outbreak constitutes a Public Health Emergency of International Concern [2]. So far, the outbreak is still on-going, the number of reported cases of 2019-nCov infection continue to increase.Several relevant literatures estimated the transmissibility of 2019-nCoV, the mean estimate of R0 for the new coronavirus pneumonia (NCP) ranges from 1.4 to 3.58[3,4]. As of 18 February 2020, NCP has caused 73400 confirmed cases including 1870 deaths, 6242 suspected cases, and 12734 cured cases in mainland released by the National Health Commission of China. Now Japan has become the second most affected country with 519 confirmed cases, and followed by Singapore with 77 confirmed cases. Person-to-person transmission of NCP has been confirmed and with several ways of transmission, moreover the asymptomatic individual is identified as a potential source of infection. Fever and cough were reported as main clinical manifestations of NCP patients[5]. Based on the clinical observations and summary, the main diagnostic criteria[6] of 2019-nCov infections are : 1) Contact history of the epidemic area and related patients; 2) Nucleic acid is positive; 3) Fever, cough, shortness of breath and other clinical symptoms; 4) Laboratory examination showed that white blood cells were normal or decreased and lymphocytes decreased; 5) Lung lesions. Especially,the 1th and 2th standards were the most important for 2019-nCov infected confirm. However, in the diagnosis and treatment process, it was found that the above diagnostic criteria could not fully meet the needs of clinical prevention and control, and some atypical clinical manifestations appeared[5,7,8], which brought great difficulties and challenges to the prevention and control of the epidemic.

Due to the large and complex population movements, the diversity of transmission pathways[5,8,9](droplets,contact, fecal-oral and aerosol pathways),and natural and intermediate hosts are not fully understood, this leads to ambiguous contact history information. In addition, some patients do not have typical symptoms of fever and cough, and there may be basic lesions or complex infections that lead to the lack of changes in the number of typical white blood cells and lymphocytes, which all increase the difficulty of clinical diagnosis. At present, nucleic acid detection was still as gold standard to confirmed NCP, but the sensitivity and specificity of NCP was not evaluated [10].According to current clinical observation, the positive rate is only 30%-50%. It has been found that patients with high clinical suspicion still have negative viral nucleic acid test for 2 times, while chest CT has displayed obvious lesions[11]. Unlucky, at the beginning, these patients failed to timely and effectively prevent and control due to negative nucleic acid test, leading to the expansion of the infection. CT diagnosis specificity is not as good as nucleic acid, but its sensitivity is significantly higher than nucleic acid. Therefore, to fight the epidemic front in Wuhan, doctor Zhang urgently called for “NPC diagnosis should not blindly believe in nucleic acid testing, chest CT findings should take as the main diagnostic basis”. Therefore, the fifth edition of NCP diagnostic criteria took chest CT as the main diagnostic basis[6]. Radiologists will encounter NCP patients with the number of patients continues to increase. Therefore, radiologists must be familiar with the CT features of NCP.

In this study, we attempt to summarize and analyze the chest CT imaging data of patients confirmed with 2019-nCov infections, and to identify the typical imaging features to improved the diagnostic accuracy of NCP, and to reduced missed diagnosis and misdiagnosis.

## 2. Materials and Methods

### 2.1 Patients and CT examination

From January 20, 2020 to February 6, 2020, 21 patients in the First people’s hospital of Foshan in Guangdong Province, China, with confirmed 2019-nCov were enrolled. these patients were divided into mild and sever group according to clinical manifestations described by the 6th clinical practice guideline of NCP in China[6]. All patients were underwent chest CT examination on an Aquilion ONE scanner (Toshiba Medical Systems;Tokyo,Japan), and imaged with 1mm slice thickness, Max 512×512. All patients were positive for 2019-nCov via laboratory testing of respiratory secretions.

Our institutional review board approved this retrospective study analyzing existing patient data with patient information de-identified and all subjects gave written informed consent. And this study involved no potential risk to patients.

### 2.2 CT Images and Clinical Data Analysis

The HRCT findings of NCP, according to previously described [9] and clinical observation, each of the 21 patients, the initial chest CT findings were categorized the predominant pattern as followed: ground glass opacification (GGO),consolidation, cobblestone/reticular pattern; mixed pattern (combination of consolidation, GGO and reticular opacities); air-bronchogram; vascular enlargement in lesion; nodule (D<30mm) and thorn pear signs; slightly dilated bronchi with thickened wall; fibrosis,etc. The the main HRCT characteristics and related exposition were showed in Fig.1. In addition, the distribution characteristics of lesions, the extent of pulmonary involvement, mediastinal lymph node status and pleural effusion were also analyzed.

**Figure 1.**
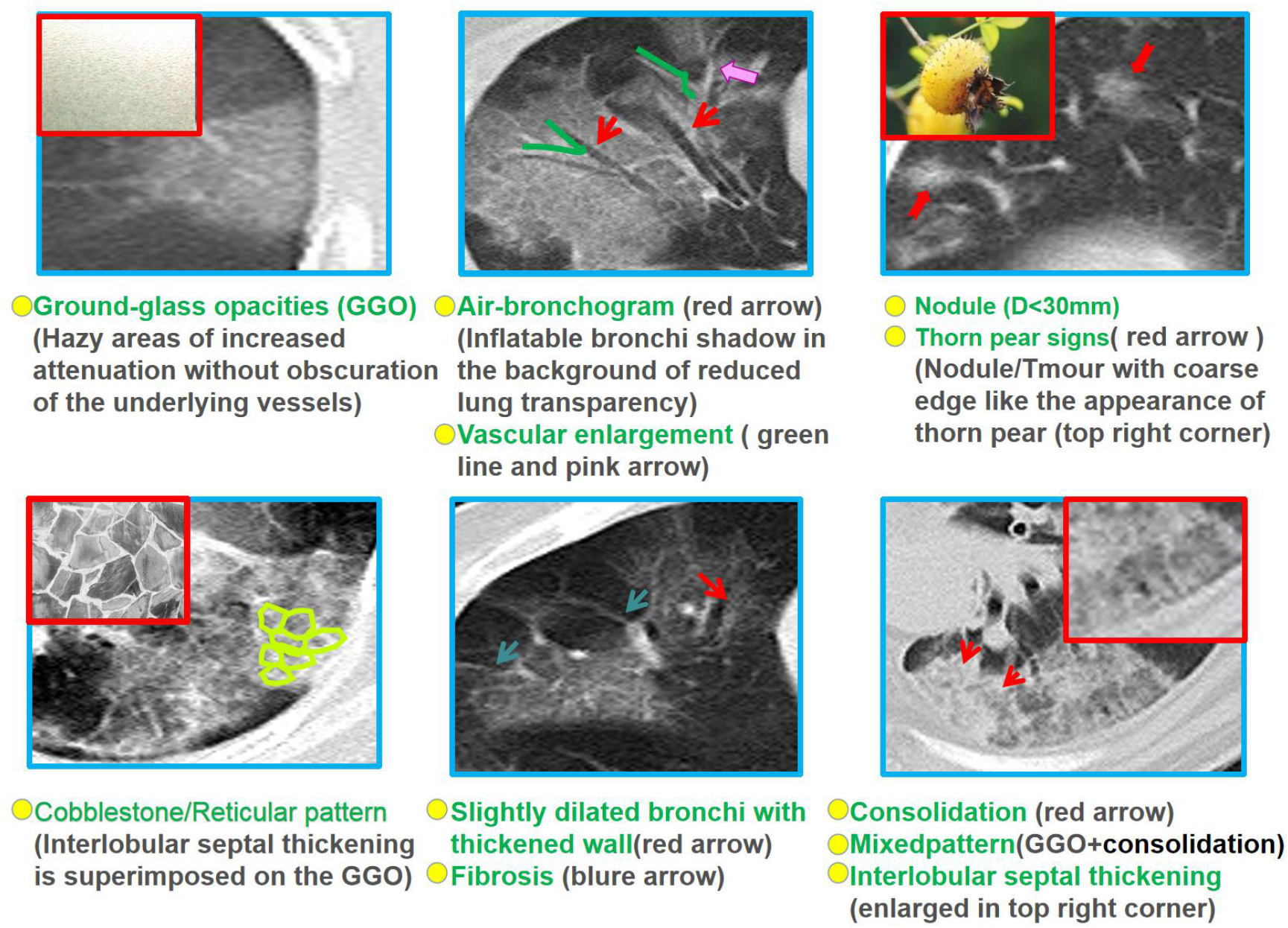
The key chest CT findings of NCP.

All CT images were reviewed on PACS by two cardiothoracic radiologists with 10 years (Dr. Xu) and 8 years (Dr.Wu) of clinical experience, respectively. Each patient’s chest CT findings was reviewed independently and final decisions reached by consensus. If the interpretations of features extracted by two primary radiologist are inconsistent, a third fellowship trained senior radiologist with 25years of experience (Dr.Pan) adjudicated a final decision.

The calinical manifestations of NCP patients, such as fever,cough, shortness of breath, weak,diarrhoea and muscle pain were analyzed.

### 2.3 Statistical Analysis

We used statistical software SPSS 2.0. The differences among the constituent ratios of various signs in NCP patients were analyzed with the X^2^ test. The differences of the constituent ratios of various signs between mild group and severe group were also analyzed with X^2^ test test. Differences of p<0.05 were considered statistically significant.

## 3. Results

### 3.1 Clinical manifestations of NCP patients

Table 1 shows the clinical characteristics of 21 patients with NCP. 21 patients all had clear contact history, including 10 men and 11 women with average age was 43.1 years. Fever (85.7%) and cough (80.9%) were the two main symptoms, followed by shortness of breath (85.7%) and weak (28.6%).Besides,diarrhoea and muscle pain were rare. Compared with patients in mild group, male proportion and onset age of patients in serve group were both higher (p<0.05). Fever and cough were found in 100% of the severe cases, but there was no significant difference between these two groups. The incidence of shortness of breath was 85.7% in the severe cases, significantly higher than 21.4% in the mild cases.

**Table 1.**
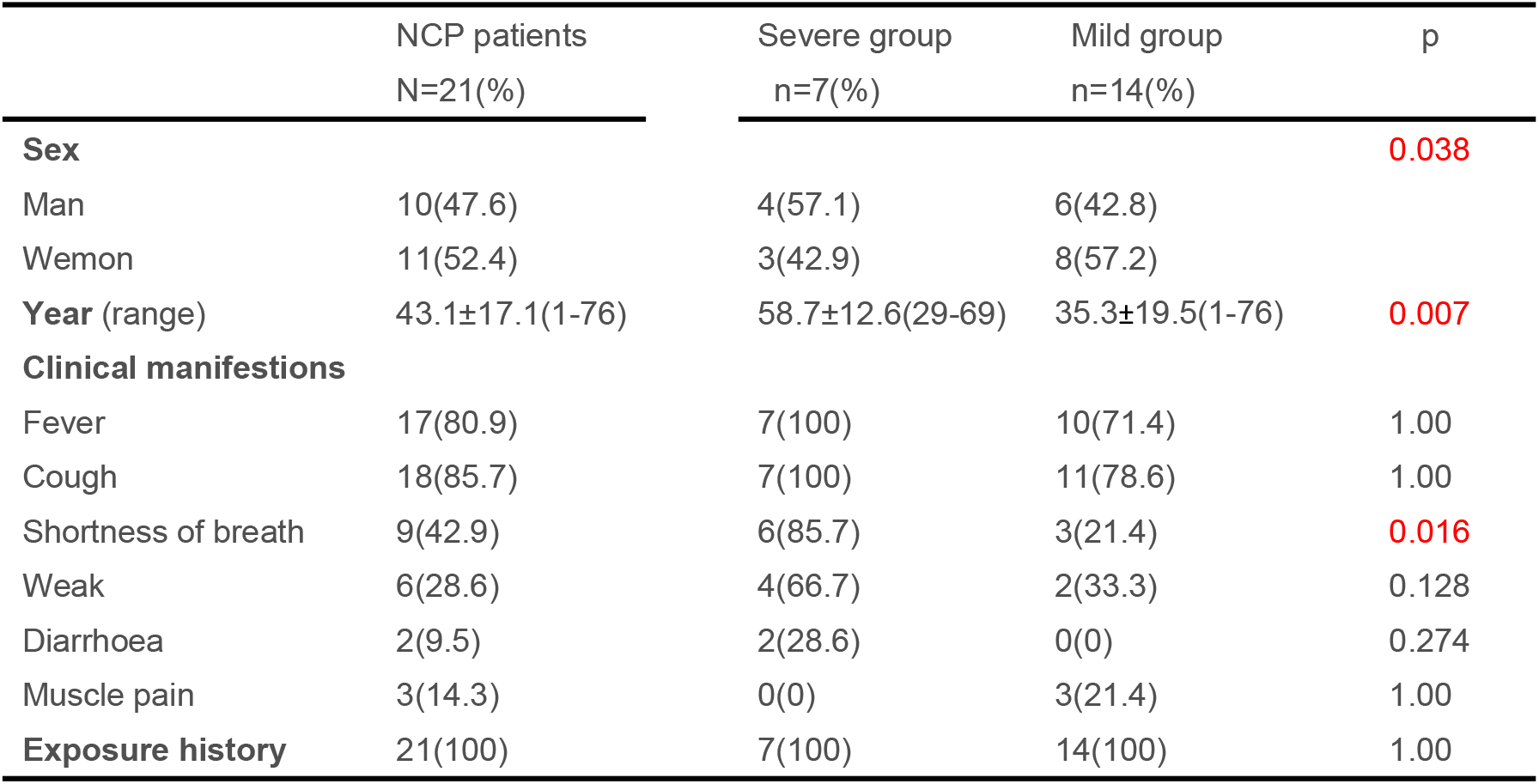
Clinical Characteristics of NCP patients

|  | NCP patients<br>N=21(%) | Severe group<br>n=7(%) | Mild group<br>n=14(%) | p |
| --- | --- | --- | --- | --- |
| **Sex** |  |  |  | **0.038** |
| Man | 10(47.6) | 4(57.1) | 6(42.8) |  |
| Wemon | 11(52.4) | 3(42.9) | 8(57.2) |  |
| **Year (range)** | 43.1±17.1(1-76) | 58.7±12.6(29-69) | 35.3±19.5(1-76) | **0.007** |
| **Clinical manifestations** |  |  |  |  |
| Fever | 17(80.9) | 7(100) | 10(71.4) | 1.00 |
| Cough | 18(85.7) | 7(100) | 11(78.6) | 1.00 |
| Shortness of breath | 9(42.9) | 6(85.7) | 3(21.4) | **0.016** |
| Weak | 6(28.6) | 4(66.7) | 2(33.3) | 0.128 |
| Diarrhoea | 2(9.5) | 2(28.6) | 0(0) | 0.274 |
| Muscle pain | 3(14.3) | 0(0) | 3(21.4) | 1.00 |
| **Exposure history** | 21(100) | 7(100) | 14(100) | 1.00 |

### 3.2 Distribution Patterns of Lesions

Lung lesions occurred in 17 (80.9%) patients with NCP, while normal lungs occurred in 4 (19.1%) patients. Among the 17 patients with pulmonary lesions, 14 (82.4%) cases showed multiple lesions in both lungs (Fig.2A). Besides,10 patients had lesions distributed in the peripheral zone (subpleural)(Fig.2A), and 7 cases showed both peripheral zone and medium-to internal zone were all involved.15 (88.2%) cases showed simultaneous involvement of multiple pulmonary lobes (≥2)(Fig.2B), among them 12 cases (70.6%) with more than 4 lobes were involved.6 patients (85.7%) in serve group displayed 4-5 lobes were involved simultaneously, which was significantly higher than that in the mild group (60%). However,there was no difference in lesion location and whether bilateral lung involvement between these two group.

**Figure 2.**
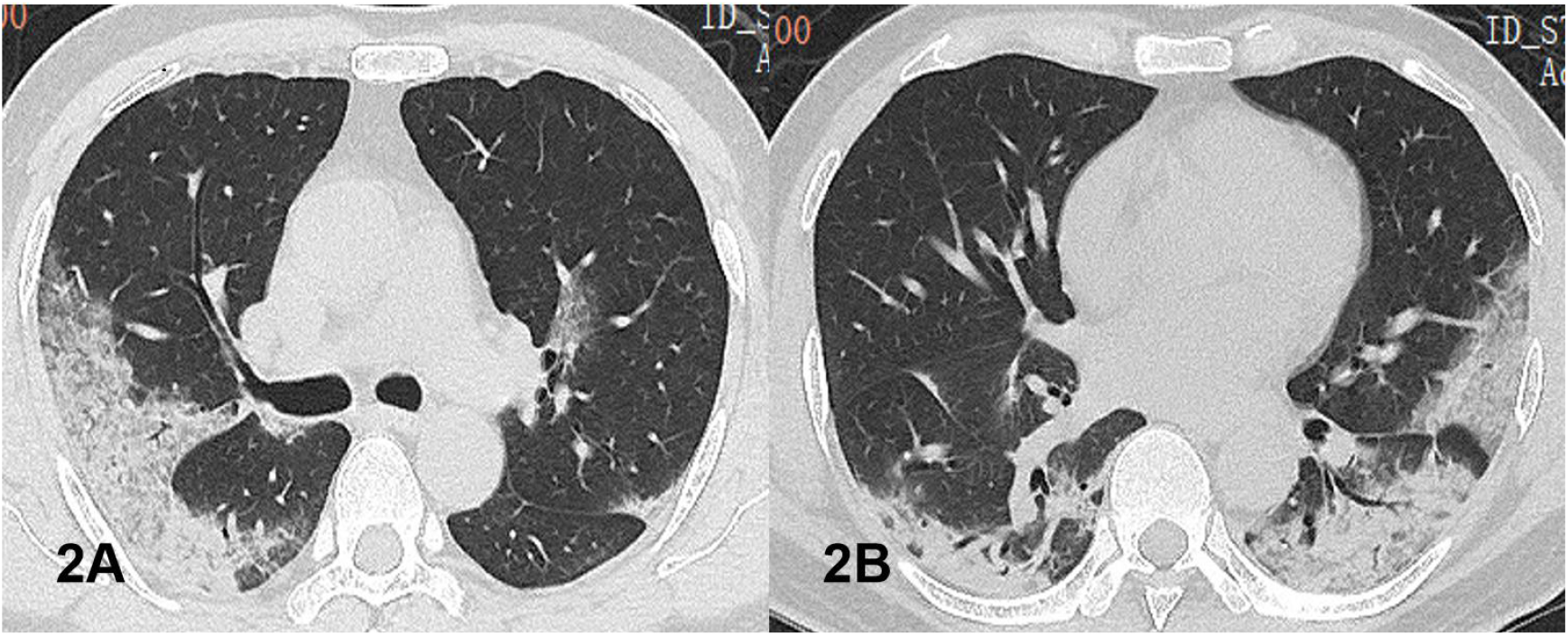
57-year old man with history of recent travel to Wuhan, presenting with fever, cough and shortness of breath. Axial thin-section non-contrast CT scan shows diffuse bilateral confluent and patchy ground-glass, air-bronchogram and consolidation, characterized by peripheral distribution. Multiple lobes were involved.

### 3.3 Representation of Major CT characteristics in NCP

Table 2 shows the manifestations of various major chest CT features in 17 patients with NCP. GGO, Vascular enlargement and Cobblestone/Reticular pattern were the three major feature of the disease(Fig.3), with an incidence of 100%, 76.5% and 70.6%, respectively and followed by Consolidation and Mixed pattern, accounting for 52.9%.In addition, mediastinal lymphadenopathy and hydrothorax had the lowest incidence of 17.6% and 5.9%, respectively.

**Table 2.** Representation of Major CT characteristics in NCP (17 Cases)

| CT Features | GGO | Con | Mix-P | Cob/Ret | Air-Bro | VEs | Fib | Nod/Tho- | Dil-Bro- | Med-Hym | Lyd |
| --- | --- | --- | --- | --- | --- | --- | --- | --- | --- | --- | --- |
| N(%) | 17(100) | 9(52.9) | 9(52.9) | 12(70.6) | 7(41.2) | 13(76.5) | 6(35.3) | 6(35.3) | 7(41.2) | 3(17.6) | 1(5.9%) |
| F | 49.741 |  |  |  |  |  |  |  |  |  |  |
| p | 0.00 |  |  |  |  |  |  |  |  |  |  |

GGO: Ground glass opacification; Con: Consolidation; Mix-P: Mixed pattern; Cob/Ret: Cobblestone/Reticular pattern; Air-Bro:Air-bronchogram; VEs:Vascular enlargement sign; Nod/Tho-: Nodule/Thorn pear signs; Dil-Bro: Dilated bronchi with thickened wall; Fib: Fibrosis; Med-Hym: Mediastinal lymphadenopathy; Hyd: Hydrothorax

**Figure 3.**
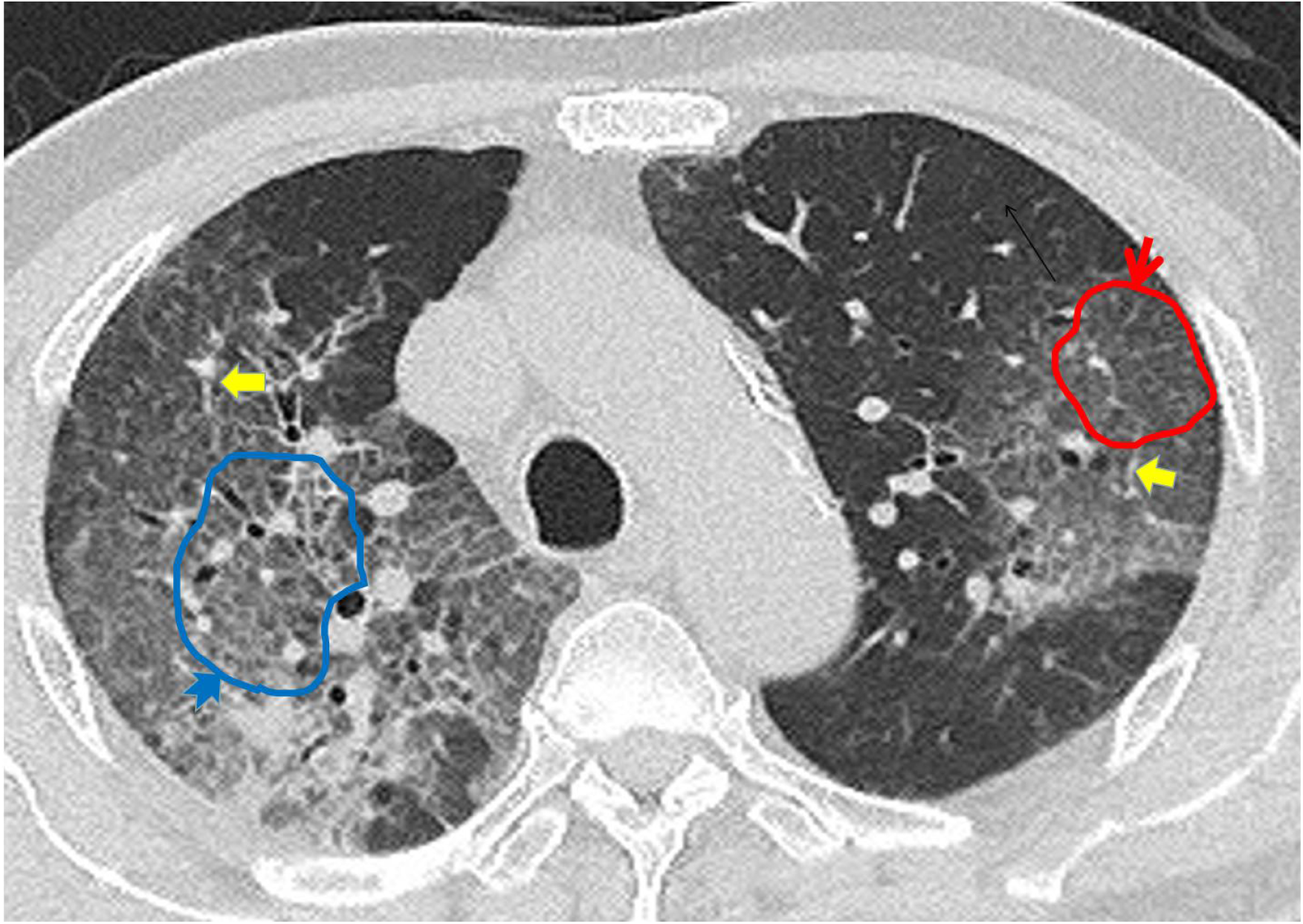
76-year old man with history of recent travel to Wuhan, presenting with fever and productive cough. Axial thin-section non-contrast CT image shows diffusion lesions in bilateral lung, mainly manifested as ground-glass opacification (red arrow and red line around region),and cobblestone/Reticular pattern(blue arrow and blue line around region). Besides,Vascular enlargement sign was observed in the lesions(yellow arrow).

### 3.4 The Differences of CT Characteristics between Mild group and Severe group

Table 3 shows the manifestations of various CT features between the two groups. GGO were present in all patients(100%) in both groups, and Cobblestone/Reticular pattern occurred in mild and sever group were 60% and 85.7%, respectively, without significant different. The consolidation, Mixed pattern and Vascular enlargement features were up to 100% in the severe group, significantly higher than 20%, 20% and 60% in the mild group. Consolidation and Mixed pattern had significant differences between the two groups (p

**Table 3.** The Differences of major CT Characteristics between two groups.

| CT Features | GGO | Con | Mix-P | Cob/Ret | Air-Bro | VEs | Fib | Nod/Tho | Dil-Bro | Med-Hym | Lyd | F | p |
| --- | --- | --- | --- | --- | --- | --- | --- | --- | --- | --- | --- | --- | --- |
| Mild group<br>n=10 (%) | 10<br>(100) | 2<br>(20) | 2<br>(20) | 6<br>(60) | 1<br>(10) | 6<br>(60) | 1<br>(10) | 5<br>(50) | 1<br>(10) | 0<br>(0) | 0<br>(0) | 48.19 | 0.00 |
| Serve group<br>n=7 (%) | 7<br>(100) | 7<br>(100) | 7<br>(100) | 6<br>(85.7) | 6<br>(85.7) | 7<br>(100) | 5<br>(71.4) | 1<br>(14.3) | 6<br>(85.7) | 3<br>(42.9) | 1<br>(14.3) | 37.80 | 0.00 |
| p | 0.425 | 0.002 | 0.002 | 0.338 | 0.004 | 0.103 | 0.035 | 0.004 | 0.035 | 0.051 | 0.415 |  |  |

=0.002). In addition, the incidence of Air-bronchogram, Dilated bronchi with thickened wall and Fibrosis in the severe group was significantly higher than that in the mild group (10%). Nodule/Thorn pear signs were found 50% in mild cases, significantly higher than those in severe cases (p<0.01). Mediastinal lymphadenopathy and hydrothorax were rare,but both occurred in severe cases. Fig.4 and Fig.5 showed the CT features of mild case and serve case underwent NCP.

**Figure 4.**
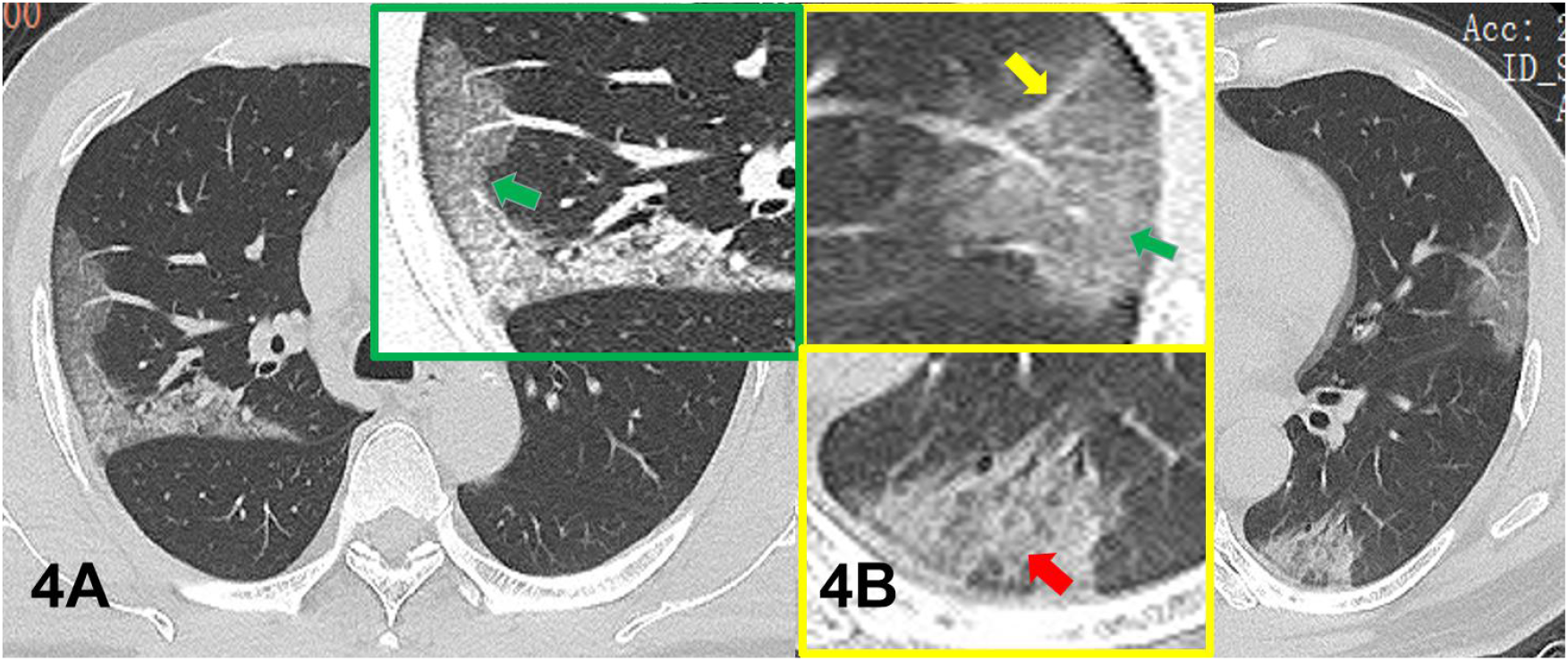
57-year old man as mild NCP patients with history of recent travel to Wuhan. Ground-glass opacities with a peripheral distribution in bilateral lung(green arrow), and with enlarged image in the upper right corner of A image. Vascular enlargement sign was observed in the lesion located in the left upper lobe, and with enlarged image in the upper left corner of B image (yellow arrow). The lesions in the left lower lobe shows cobblestone/reticular pattern, and with enlarged image in the lower right corner of B image(red yellow).

**Figure 5.**
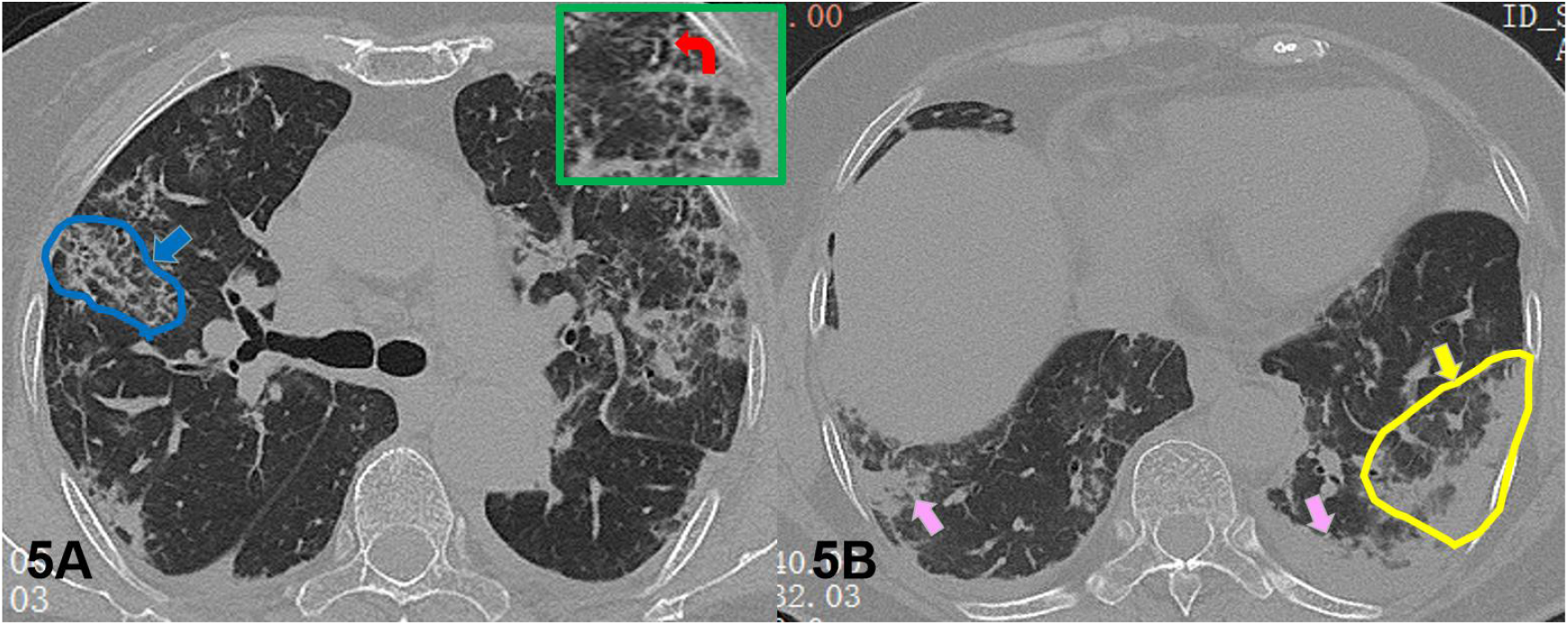
68-year old female as serve NCP patients with history of recent travel to Wuhan, presenting with fever and productive cough. Axial thin-section non-contrast CT image shows diffusion lesions in bilateral lung. A showed many CT features such as ground-glass opacification,cobblestone/reticular pattern(blue arrow and bule line around region), frbrosis and dilated bronchi with thickened wall (the enlarged image in the upper right corner, red arrow). B showed consolidation (pink arrow) and mixed pattern (yellow arrow and yellow line around region) in the bilateral lower lobes.

## 4. Discussion

### 4.1 Clinical features of NCP patients

All 21 patients in our study all had a history of recent travel to Wuhan or directly contact with NCP patients.This results was convincing proof that the disease is characterized by human-to-human transmission. A new study revealed that the reproductive number of COVID-19 is up to 3.28 [3]. However, in this study, the patients did not be acquired a clear transmission route, because the virus may have multiple transmission modes[12,13]. Previous study demonstrated that common symptoms at onset of the illness were fever, cough and shortness of breath and with less common symptoms were sputum production, headache and diarrhoea etc[4,5,7,12,13]. Our results revealed that main symptom of NCP patients manifested as fever (85.7%), cough (80.9%), followed by shortness of breath(42.9%)and fatigue(28.6%),diarrhea and muscle pain,which was consistent with previous studies.However,a large clinical cases(1099 cases) study by professor zhong et al [7] found that only 43.8% showed fever in the early stage, but up to 87.9% showed fever after hospitalization.It dmonstrated that fever is closely related to the development of the disease, therefore fever as a screening indicator may miss a large number of early patients. In addition,our study and all previous showed that diarrhea was rare, and we found that diarrhea only occurred in serve patients. Therefore, it is necessary to pay attention to the damage of digestive tract function in these patients and be alert to the risk of fecal-oral infection. 85.7% of severe cases manifested shortness of breath, were significant higher than that of mild cases. Shortness of breath would be a better indicators to reflect the disease’s progression. A new study[12] base on 72314 patients revealed that middle-aged(30-79 years old) and elderly men (51.4%) were prone to the NCP,and looking at the sex ratio, men are more likely to die (2.8%) than women (1.7%). our results showed NCP patients with average 43.1 years old, and men are more likely to be worse(57.1%), showed similar characteristics.

### 4.2. CT features of NCP and the relationship with the disease development

Lung lesions in NCP patients mainly manifested as subpleural multiple lesions in bilateral lung[14,15],our results showed 82.4% cases displayed multiple lesions in bilateral lungs, and all cases showed significant peripheral distribution characteristics, wich was consistent with previous studies[14-16].We speculated that the main reasons for the distribution characteristics of NCP as followed: 1)The 2019-nCov infect alveolar epithelial cells through angiotensin-converting enzyme 2 (ACE2) and cause acute lung injury[17].The peripheral lobe of the lung is relatively well developed, abundant alveolar epithelial cells may express more ACE2. 2) The ventilation function of alveolar tissue in the peripheral part of the lung is relatively weak, which may more easily lead to the aggregation of high concentration of virus particles and the onset of disease. In addition, some scholars believe that virus particles are extremely small (2019-nCov is about 120nm), which can directly enter the distal alveoli with respiration. In this study, CT features of NCP patients mainly displayed GGO (100%), vascular enlargement sign (76.5%) and cobblestone/reticular pattern (70.6%), as followed by consolidation and mixed pattern (GGO and consolidation) with about 52.9%. While mediastinal lymph nodes and pleural effusion were rare, which was basically consistent with literature reports[14,15].In general, the lung characteristics of NCP patients were mainly associated with pulmonary interstitial inflammation, which was similar to Middle East respiratory syndrome(MERS)[18] and Severe Acute Respiratory Syndrome(SARS)[19] caused by coronavirus. In fact, the pathological characteristics of NCP were confirmed to be similar to SARS and MERS[20]

Previous reports revealed that GGO as the key CT feature.It can exists for a long time throughout the course of the disease, and may be unique CT feature in some early patients[15,16]. In the recent study, GGO feature was observed in all NCP patients. Pathological basis of GGO is mainly diffuse alveolar injury, accompanied by cell fibromyxoid exudate,and lung transparent membrane formation. GGO at the early stage could be completely absorbed and improved after effective treatment [21]. With the development of the disease, the interlobular septa thickened on the basis of GGO, and showed typical cobblestone/reticular pattern,and when increased exudation in the alveoli and alveolar collapse resulted in consolidation. Compared with the mild group patients, the incidence of consolidation, mixed pattern, air-bronchogram, dilated bronchi with thickened wall and fibrosis significantly increased in severe patients. In addition, cobblestone/reticular pattern and vascular enlargement sign were both common findings in severe patients without significant different. It is suggested that the changes of pulmonary imaging characteristics can effectively reflect the changes of lung tissue and functional injury.Vascular enlargement in or around the lesions were considered to be an important features for NCP diagnosis,76.5% patients in our study presented this features. This may be due to inflammatory vascular congestion, or to thickening or fluid buildup in the perivascular interstitium[20]. Additionally,all these findings suggest the presence of fibrosis, which also presented commonly in patients underwent SARS and MERS.In this study, consolidation and mixed manifestations were observed in 52.9% of NCP patients, and the incidence rate was up to 100% in severe patients. Combined with previous literature results, lung tissue appeared multiple consolidation with the aggravation of the disease. That’s the main difference from MERS[18]. 6 patients (85.7%) in serve group displayed 4-5 lobes were involved simultaneously, which was significantly higher than that in the mild group (60%).Our result demonstrated that almost all lobes involvement exists in severe patients, which well explains the pathologic basis for the high incidence of dyspepsia. It was found that CT can not only detect early and diagnose NCP accurately, but also follow up the change trend of CT features can effectively predict the outcome of NCP patients.

### 4.2 Limitations

The main limitations of this study, firstly, the small number of NCP cases enrolled in this study and additional study cases are needed in the following study. Besides, This study lacks patient follow-up data, so further follow-up is needed to better observe the relationship between CT signs and clinical course.

## 5. Conclusions

Fever and cough are the main clinical features of NCP. CT showed multiple peripheral lesions in bilateral lungs, mainly characterized by GGO, paving stones and vascular thickening. There were some differences in CT manifestations between mild cases and severe cases. In severe cases, pulmonary consolidation, mixed consolidation, slight bronchial dilatation and thickening, bronchial gas image and fibrosis were significant, which were consistent with NCP as pathological manifestations of pulmonary interstitial lesions. According to the characteristics of chest CT, early accurate diagnosis and prognosis of NCP can be achieved.

## Data Availability

All data are from the First people's hospital of Foshan, the data is available.

## Abbreviations

2019-nCov: 2019 Novel Coronavirus
NCP: new coronavirus pneumonia
SARS: Severe Acute Respiratory Syndrome
MERS: Middle East Respiratory Syndrome
GGO: Ground glass opacification
Con: Consolidation
Mix-P: Mixed pattern
Cob/Ret: Cobblestone/Reticular pattern
Air-Bro: Air-bronchogram
VEs: Vascular enlargement sign
Nod/Tho-: Nodule/Thorn pear signs
Dil-Bro: Dilated bronchi with thickened wall
Med-Hym: Mediastinal lymphadenopathy
Hyd: Hydrothorax

## Acknowledgements

This study was supported by Y J Ye,department of radiology,the first people’s Hospital of Foshan affiliated to sun yat-sen university,Foshan,China.

## Declaration of interests

We declare no competing interests.

## Notes

### Competing Interest Statement

The authors have declared no competing interest.

